# Inconsistency of AI in Intracranial Aneurysm Detection with Varying Dose and Image Reconstruction

**DOI:** 10.1101/2024.11.26.24317954

**Authors:** Leonie Goelz, Angelo Laudani, Ulrich Genske, Michael Scheel, Georg Bohner, Hans-Christian Bauknecht, Sven Mutze, Bernd Hamm, Paul Jahnke

## Abstract

**Background:** Scanner-related changes in data quality are common in medical imaging, yet monitoring their impact on diagnostic AI performance remains challenging.

**Purpose:** To assess the consistency of an FDA-approved AI solution for intracranial aneurysm detection across changes in image data quality caused by dose and image reconstruction.

**Methods:** Consistency testing was performed using a head CT phantom designed for AI evaluation, replicating a patient with three intracranial aneurysms in the anterior communicating artery (ACoA), middle cerebral artery (MCA), and basilar artery (BA). The phantom was examined three times at 21 doses ranging from 0.47 to 20.09 mGy using iterative reconstruction (IR) and filtered back projection (FBP). Aneurysm labeling by an AI solution approved for triage and notification of intracranial aneurysms was analyzed. Five neuroradiologists evaluated all examinations for aneurysm visibility.

**Results:** The AI solution labeled 74.6% of ACoA, 92.9% of MCA, and 2.4% of BA aneurysms, while reader experiments yielded aneurysm visibility rates of 98.6%, 99.8%, and 95.4%, respectively. The AI demonstrated stable performance within the medium dose range but produced inconsistent results at doses below 8 mGy with IR, 7 mGy with FBP, and also above 14 mGy with FBP. Doses below 1 mGy with IR and 2 mGy with FBP led to a complete lack of AI response to any aneurysm. Readers noted at least one visible aneurysm in every case and reported 100% visibility for all aneurysms at doses above 2 mGy.

**Conclusions:** Diagnostic AI shows inconsistent performance when image data quality is not optimal and requires more stringent quality standards than radiologists. Standardized consistency testing reveals AI performance issues when these quality requirements are not achieved.

## Introduction

Intracranial aneurysms (IAs) affect approximately 2-3% of the global population^1-3^. While many remain asymptomatic, rupture can lead to devastating consequences such as disablement or death. Computed tomography angiography (CTA) plays a key role in diagnosing IA due to its speed, availability, non-invasiveness, and diagnostic accuracy^4,5^. While traditionally interpreted by radiologists, artificial intelligence (AI)-enabled software tools show promise in enhancing aneurysm detection and management^6,7^.

The U.S. Food and Drug Administration (FDA) has approved several AI-enabled applications for managing IA based on CTA. However, clearance of these applications does not guarantee reliable model performance in actual clinical use^8^. Variations in CT scanners, protocols, and reconstruction methods in specific clinical practice may differ from those during development, implementation, and validation of AI models, potentially affecting data quality. It remains uncertain whether these applications can reliably perform on such varied data types.

Current methods for independent AI validation involve publicly released datasets as benchmarks to assess algorithm performance^9,10^. These initiatives enable evaluation outside the developmental environment, offering an independent estimate of performance and generalizability^11^. However, this information does not indicate how the algorithm will perform in a specific radiology facility. Furthermore, if imaging systems or protocols are modified, it remains uncertain whether the AI will still perform as expected^12^. Even without apparent modifications, the performance of AI algorithms is prone to decay and benefits from regular monitoring^13^. Addressing these issues requires iterative local validation steps in the specific imaging department^14^. However, each of these iterations requires the collection of patient data, which is time-consuming and impractical for efficiently validating and optimizing scanners, data quality, and AI functionality.

In contrast, phantoms featuring human tissue and pathology patterns enable controlled access to scan data and provide opportunities for standardized consistency testing of AI. In this study, a phantom simulating a patient with three IAs was used to evaluate a diagnostic AI algorithm for intracranial aneurysm detection. We hypothesized that variations in data quality can influence AI output, and that such variations may have a different impact on AI than on image interpretation by neuroradiologists. Therefore, the aim of our study was to assess the consistency of an FDA-approved AI solution for IA detection across changes in image data quality caused by dose variations and image reconstruction.

## Methods

### Study design

The study was conducted prospectively following ethics approval. A CT phantom simulating a patient with three intracranial aneurysms was examined using 21 doses and two reconstruction algorithms. Aneurysm detection was evaluated using a commercial AI solution and by five neuroradiologists.

### Phantom

A realistic head phantom (50-03, PhantomX, Berlin, Germany) specifically designed for AI evaluation was employed in this study. The phantom simulated a patient post contrast medium injection in arterial phase and featured three intracranial aneurysms of the anterior communicating artery (ACoA), middle cerebral artery (MCA), and basilar artery (BA) with maximum diameters of 4 mm, 4 mm, and 2 mm, respectively. It was fabricated using a technology based on previously published methods^15,16^. For image acquisition, the phantom was positioned within a support pillow in the isocenter of the CT scanner gantry reflecting the typical positioning during clinical routine. Figure 1 shows the phantom setup and CT images of the phantom visualizing the three aneurysms.

**Fig. 1:**
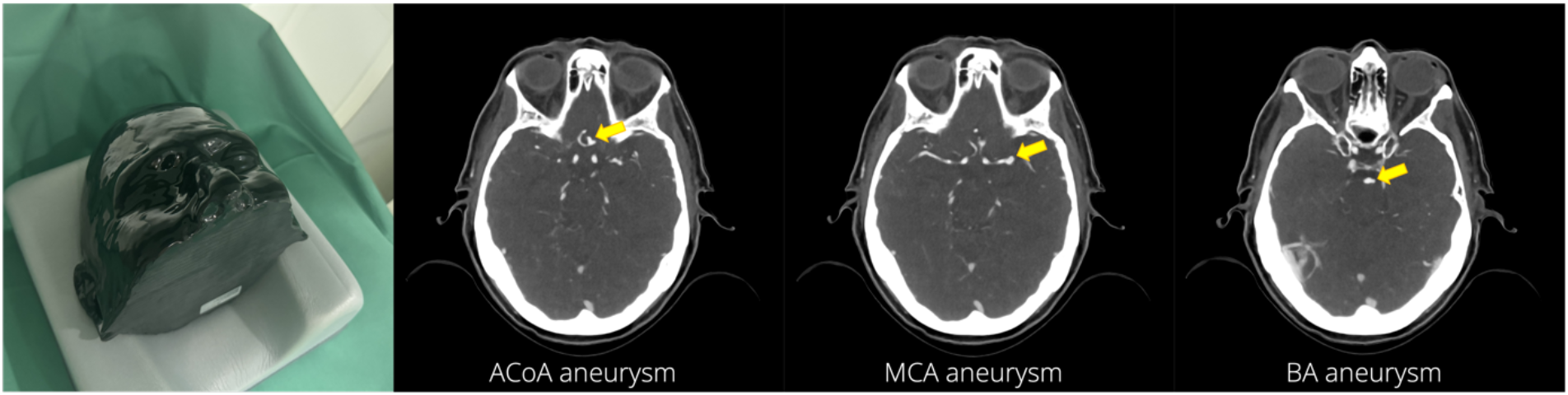
Setup and CT images of the phantom. Photograph shows the phantom positioned in the CT scanner for image acquisition. CT images of the phantom visualize the three aneurysms in the anterior communicating artery (ACoA), middle cerebral artery (MCA), and basilar artery (BA).

### Image acquisition

Images were obtained using a 128-row CT scanner (Somatom AS, Siemens Healthineers, Erlangen, Germany). The tube voltage was 120 kVp, the pitch was 0.9, the image matrix was 512 x 512 pixels and the pixel size ranged between 0.39 and 0.42 mm (median: 0.41 mm). Twenty-one computed tomography dose indices (CTDI) ranging from 0.47 to 20.09 mGy were employed. To this end, the tube current and rotation time were varied as detailed in table 1. Images were reconstructed using the department’s standard iterative reconstruction (IR) algorithm Sinogram Affirmed Iterative Reconstruction (SAFIRE, I26f kernel) and filtered back projection (FBP, B26f kernel). Three acquisitions were performed per dose and image reconstruction, totaling 126 scans. Images were reconstructed in axial orientation with 0.75 mm slice thickness.

**Table 1:** Computed tomography dose indices (CTDIvol), rotation times, and tube current-time products used for image acquisition.

| **CTDIvol (mGy)** | **Rotation time (s)** | **Tube current-time product (mAs)** |
| --- | --- | --- |
| 0.47 | 0.3 | 7 |
| 1.01 | 0.5 | 15 |
| 2.02 | 0.5 | 30 |
| 3.00 | 0.5 | 44 |
| 4.01 | 0.5 | 59 |
| 5.02 | 0.5 | 74 |
| 6.03 | 0.5 | 89 |
| 7.05 | 0.5 | 104 |
| 8.06 | 0.5 | 119 |
| 9.07 | 0.5 | 134 |
| 10.08 | 0.5 | 149 |
| 11.09 | 0.5 | 164 |
| 12.11 | 0.5 | 179 |
| 13.04 | 0.5 | 193 |
| 14.06 | 0.5 | 208 |
| 15.07 | 0.5 | 223 |
| 16.04 | 0.5 | 238 |
| 17.05 | 0.5 | 253 |
| 18.07 | 0.5 | 268 |
| 19.08 | 0.5 | 283 |
| 20.09 | 0.5 | 298 |

### AI image assessment

The study site had a commercial, cloud-based AI solution (AIDOC, Tel Aviv, Israel) integrated into the clinical workflow, which included algorithms for detection of IA and large vessel occlusion in CTA, and intracranial hemorrhage in non-contrast head CT. The solution was set up so that CTAs were automatically processed starting with their export to a virtual machine within the department. Here, the software’s orchestrator identified the examined body part and study protocol to ensure processing by the appropriate algorithms. In addition, the orchestrator conducted an initial check for technical issues rendering scans unsuitable for analysis, such as excessive motion artifacts, metal artifacts, inadequate field of view, or insufficient arterial contrast. When the initial check was passed, CTAs with positive IA findings were highlighted in the cloud for prioritization, and all results (positive or negative) were fed back into the picture archiving and communication system as color-coded heatmaps or negative summary reports. The heatmaps and summary reports resulting from the phantom examinations were independently reviewed by two board-certified radiologists with 10 years of experience to determine positive and negative aneurysm labels, followed by a consensus reading.

### Reader image assessment

Five board-certified neuroradiologists participated in a reading experiment to evaluate aneurysm visibility and image quality. Their experience ranged from 9 to 24 years (median: 17 years). Each reader was presented with all 126 datasets in random order and asked to indicate the visibility of each of the three aneurysms (1=yes, 2=no). In addition, readers were asked to evaluate image quality for the assessment of each aneurysm (3=good, 2=intermediate, 1=poor). No consensus reading was done. Readers were familiarized with the experimental setup in a training session preceding the reading experiment. All readings were performed using the open-source platform *Human Observer Net*^17^.

### Data analysis

True positive aneurysm labels are presented in absolute numbers. For the analysis of label sizes and intensities, aneurysm labels had to be segmented. To this end, a pixel value of 100 in the blue channel of the 8-bit RGB (red, green, blue) color-coded heatmaps was defined as the maximum threshold for positive aneurysm labels. Label sizes were calculated by multiplying the number of positive pixels by the pixel size. For the analysis of pixel intensities, RGB values of labels were converted into grayscale values, inversed, normalized, and then averaged. Average label sizes and intensities across three repeated acquisitions were compared between aneurysms using the Mann-Whitney U-test and Bonferroni correction to adjust p-values for multiple comparisons. Reproducibility was analyzed using the coefficient of variation (CV), employing only results from those doses and reconstructions with three positive labels. Aneurysms rated as visible by readers are presented in absolute numbers. Image quality ratings of 1 (poor) were assigned the value 0 if readers also rated the aneurysm as not visible. Image quality ratings were averaged per aneurysm and reader across three repeated CT examinations per dose and image reconstruction method. Mean values of all five readers were used to compare image quality ratings between aneurysms and reconstruction methods using the Mann-Whitney U-test with Bonferroni correction of p-values. Differences were interpreted as significant for p < 0.05.

## Results

### Imaging and AI output

The orchestrator processed all scans, AI results (positive findings on heatmaps or negative summary reports) were obtained without exception. Out of the 126 scans, 117 (92.9%) generated heatmaps that implied true positive triage decisions, while 9 (7.1%) resulted in negative outputs corresponding to false negative triage decisions. Figure 2 shows examples of phantom scans and AI output.

**Fig. 2:**
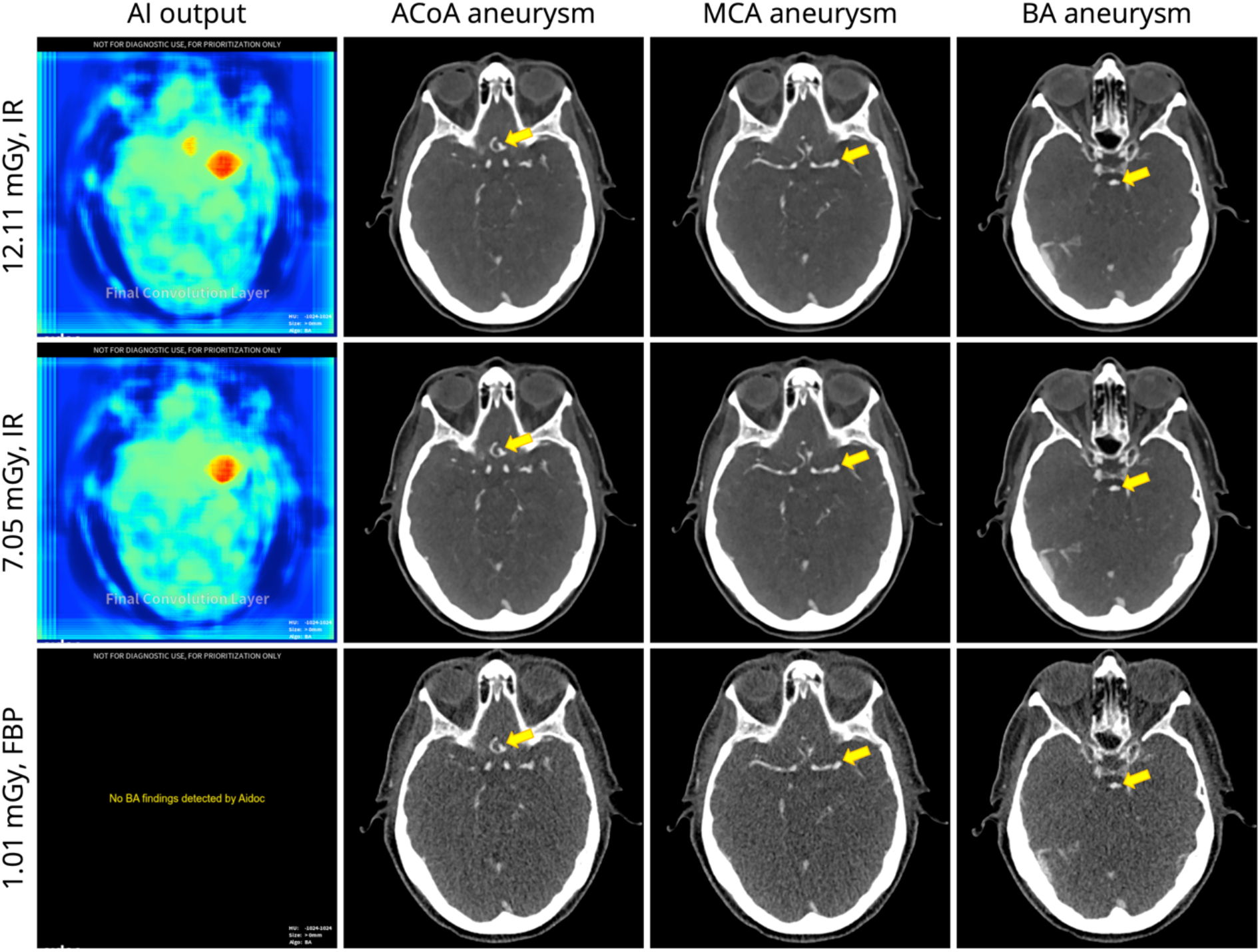
Examples of AI output and corresponding CT images. First row: The AI-generated heatmap displays true positive labels for aneurysms in the anterior communicating artery (ACoA) and middle cerebral artery (MCA), but not for the basilar artery (BA). Second row: The heatmap indicates a true positive label for the MCA aneurysm, but not for the ACoA and BA aneurysms. Third row: A false negative summary report generated by the AI solution, where BA stands for brain aneurysm. IR = iterative reconstruction, FBP = filtered back projection.

### AI labeling of the MCA, ACoA, and BA aneurysms

Overall, the AI solution generated 214 heat map labels. Review of the labels by two radiologists yielded no false positive intracranial aneurysm finding and no disagreement between the two readers. Figure 3 presents true positive aneurysm labels per aneurysm. Label sizes and intensities are detailed in figure 4. Across all 126 acquisitions, the MCA aneurysm was most frequently labeled (117 labels, 92.9%), followed by the ACoA aneurysm (94 labels, 74.6%) and the BA aneurysm (3 labels, 2.4%). Also, the size and intensity of MCA labels were greater than those of ACoA and BA labels (p < 0.001) and they were associated with better reproducibility. CVs for label size were 0.08 (MCA) and 0.25 (ACoA) with IR and 0.05 (MCA) and 0.26 (ACoA) with FBP. CVs for label intensity were 0.04 (MCA) and 0.08 (ACoA) with IR and 0.05 (MCA) and 0.08 (ACoA) with FBP.

**Fig. 3:**
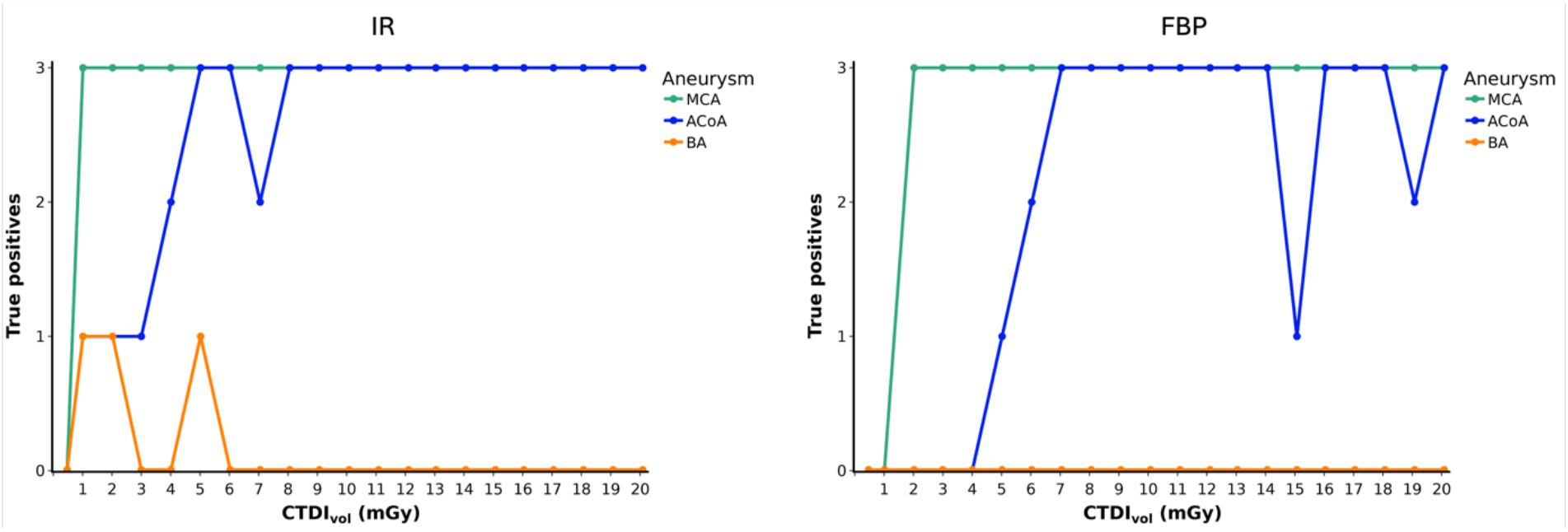
True positive aneurysm labels in heatmaps generated by the AI solution. Graphs display the absolute numbers of true positive aneurysm labels across three repeated CT examinations per dose and image reconstruction method. IR = iterative reconstruction, FBP = filtered back projection, MCA = middle cerebral artery, ACoA = anterior communicating artery, BA = basilar artery, CTDI_vol_ = computed tomography dose index.

**Fig. 4:**
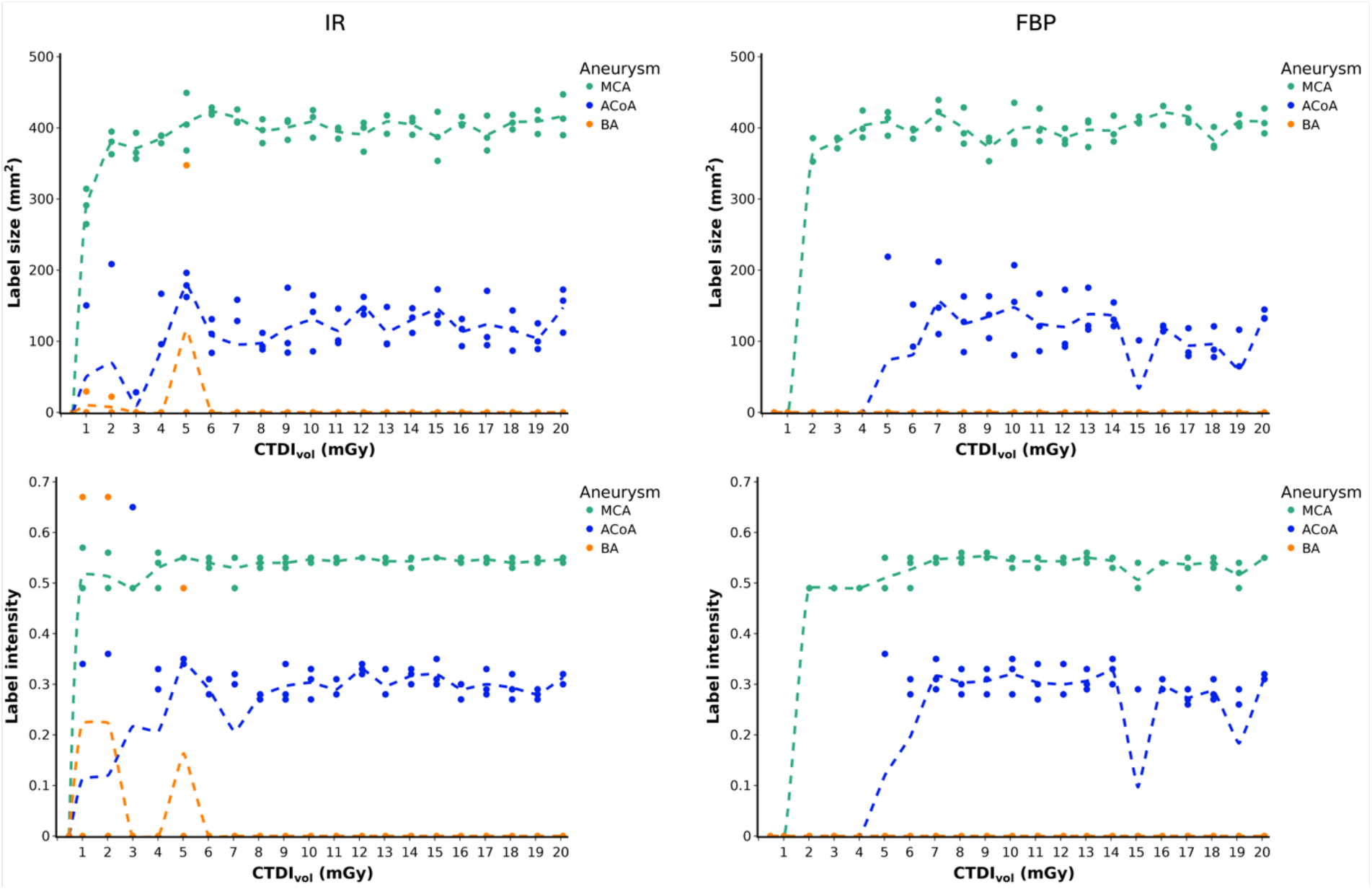
Sizes and Intensities of aneurysm labels. Graphs present the size (upper row) and intensity (lower row) of aneurysm labels generated by the AI solution in three repeated CT examinations per dose and image reconstruction method. Dotted lines indicate mean values. IR = iterative reconstruction, FBP = filtered back projection, MCA = middle cerebral artery, ACoA = anterior communicating artery, BA = basilar artery, CTDI_vol_ = computed tomography dose index.

### AI aneurysm labeling in IR-reconstructed acquisitions

Using the department’s standard image reconstruction algorithm, IR, doses of ≥8 mGy resulted in correct and reproducible labeling of the MCA and ACoA aneurysms. In contrast, AcoA aneurysm labels were not reproducible at doses below 8 mGy, except at 5 and 6 mGy. Interestingly, the only acquisition that produced true positive labels for all three aneurysms present in the phantom occurred at 5 mGy with IR. However, this result was non-reproducible as the BA aneurysm was labeled in only one of three repeated acquisitions. Incidental BA labels were also produced at 1 and 2 mGy, but these labels were small and slightly offset when superimposed with the original scan data (Fig. 5). The lowest dose of 0.5 mGy produced no aneurysm labels, while the MCA label was robust at higher doses. False positive vessel occlusion alerts were generated at doses of 3 mGy in 1 of 3 repetitions and 2 mGy in 2 of 3 repetitions.

**Fig. 5:**
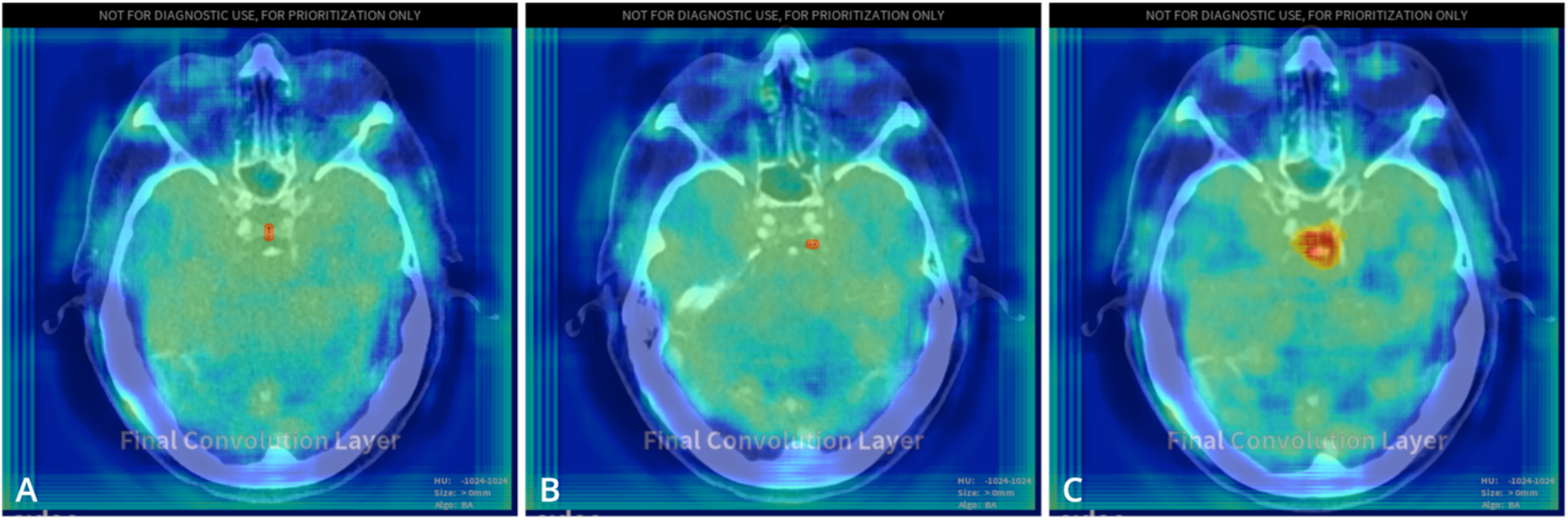
Heatmaps generated by the AI solution with positive labels for the basilar artery aneurysm superimposed on corresponding CT images. Labels in CT examinations at 1.01 mGy (A) and 2.02 mGy (B) are slightly offset. (C) The label in a CT examination at 5.02 mGy aligns with the aneurysm. All CT examinations presented were reconstructed using iterative reconstruction.

### AI aneurysm labeling in FBP-reconstructed acquisitions

Using FBP for image reconstruction resulted in decreased AI labeling of the MCA (57 vs. 60 with IR), ACoA (42 vs. 52 with IR), and BA (0 vs. 3 with IR) aneurysms. ACoA aneurysm labeling deteriorated at doses below 7 mGy, similar to IR. However, unlike with IR, ACoA labeling was also non-reproducible at higher doses of 15 and 19 mGy. None of the aneurysms were labeled at 0.5 mGy, as with IR, and additionally at 1 mGy. A false positive vessel occlusion alert was generated at 2 mGy in 1 of 3 repetitions.

### Reader assessment

Figure 6 presents the results of the reading experiment. Numerical results are provided in supplementary tables 1 and 2. Readers found at least one aneurysm visible in 100% of cases.

**Fig. 6:**
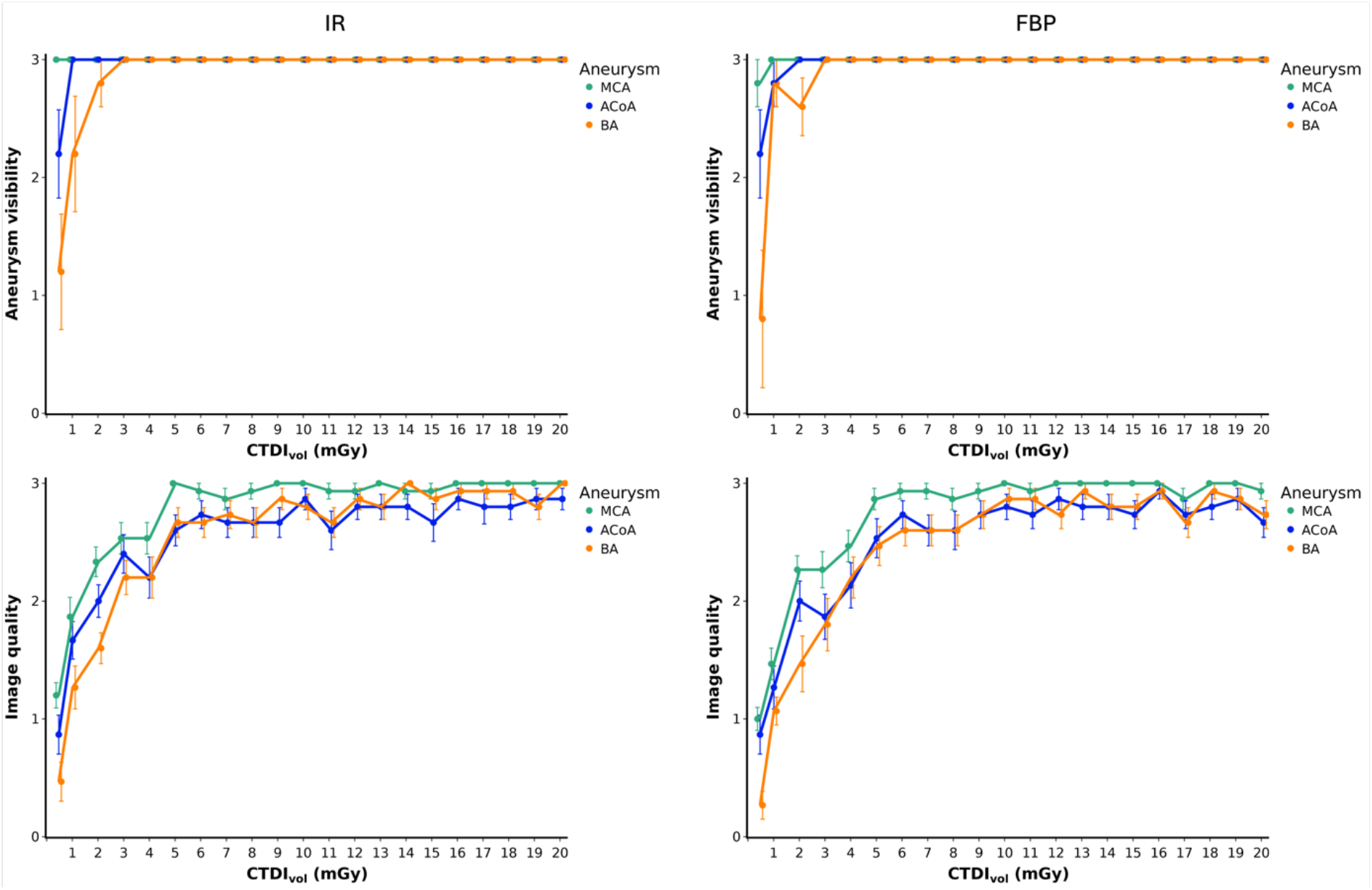
Results of aneurysm assessment by five neuroradiologists. Upper row: Absolute number of CT examinations in which aneurysms were rated as visible across three repeated CT examinations per dose and image reconstruction method. Lower row: Image quality ratings per aneurysm, dose, and image reconstruction method. Mean results of the five readers are presented; error bars indicate standard errors of the mean. IR = iterative reconstruction, FBP = filtered back projection, MCA = middle cerebral artery, ACoA = anterior communicating artery, BA = basilar artery, CTDI_vol_ = computed tomography dose index.

With the exception of one reader and one FBP-reconstructed scan at 0.47 mGy, readers always found the MCA aneurysm visible (629/630 positive responses, 99.8%). The ACoA aneurysm was rated to be visible in 98.6% of cases (621/630), and the BA aneurysm in 95.4% of cases (601/630). One hundred percent of aneurysms were found to be visible at doses >2 mGy, regardless of image reconstruction, while doses ≤2 mGy degraded aneurysm visibility in both IR- and FBP-reconstructed images. Image quality ratings deteriorated at lower doses in both IR and FBP reconstructions. Image quality ratings for the MCA aneurysm were superior to those for ACoA and BA aneurysms in both IR (p ≤ 0.038) and FBP (p ≤ 0.012), whereas no difference was found between the ACoA and BA aneurysms regardless of the image reconstruction method (p ≥ 0.893). There was also no difference in image quality ratings between the two reconstruction methods for any of the three aneurysms (p = 1).

## Discussion

Scanner-related changes in data quality are common in medical imaging, yet monitoring their impact on diagnostic AI performance remains challenging. In this study, we performed standardized consistency testing of a commercial AI solution for intracranial aneurysm detection across changes in CT data quality caused by dose and image reconstruction. The assessment was based on a phantom replicating a patient with aneurysms in the anterior communicating artery (ACoA), middle cerebral artery (MCA), and basilar artery (BA). Our findings reveal differences between aneurysm types, with only 3 BA labels (2.4%) versus 94 (74.6%) for the ACoA and 117 (92.9%) for the MCA. The AI generated inconsistent results at doses below 8 mGy with iterative reconstruction and at doses below 7 mGy and above 14 mGy with filtered back projection (FBP). Neuroradiologists were less affected by variations in data quality and reported 100% visibility of all three aneurysms at doses above 2 mGy.

Diagnostic AI solutions are developed and approved using datasets that are representative of the target population to ensure adequate generalizability for clinical application^18^. Naturally, these datasets tend to include more common conditions, while rarer cases are less represented. BA aneurysms are relatively rare, and very small aneurysms pose a particular challenge for AI, which explains why the BA aneurysm generated almost no response in our study^19^. Additionally, doses below and above the typical range used in most institutions are less represented in AI development, resulting in inconsistent or negative AI responses. This issue affected the ACoA aneurysm more than the MCA aneurysm due to its more challenging location at the base of the skull, which resulted in lower network attention.

AI users are often unaware of the limitations a particular algorithm may have in their specific setting, and changes to equipment or imaging protocols can lead to unexpected AI performance^20^. For instance, a simple software update to mammography systems was reported to cause unexpected AI behavior, significantly increasing recall rates in breast cancer screening^12^. Our findings demonstrate similar unexpected AI behavior, with inconsistent responses triggered by dose increase with FBP, which contrasts with the conventional expectation that higher doses generally improve diagnostic image quality for radiologists.

Our results do not contradict studies reporting the potential of AI to outperform radiologists in aneurysm detection^21^. However, they reveal substantial differences in susceptibility to changes in data quality between radiologists and AI. This has implications for radiological practice as radiologists cannot adapt imaging technologies solely based on their own needs if they intend to use AI. Instead, they must carefully revalidate whether changes also lead to satisfactory AI functionality^20^. This requires consistency testing, which is also highly relevant beyond intended modifications of imaging equipment to monitor and detect AI decay^13,20^.

Current consistency test concepts rely on monitoring AI during clinical routine, meaning inconsistent AI performance becomes apparent if the AI output does not align with radiologists’ assessments^22^. This process is resource-intensive, lacks objectivity and standardization, and its retrospective nature potentially jeopardizes patients exposed to inadequate AI performance. Phantoms are standard instruments for prospective consistency testing but have not been applicable for AI due to their lack of anatomical and pathological features. Dedicated phantoms with realistic features have therefore been defined as a key element for validation and quality control in imaging and AI ecosystems^23^. In this study, we employed such a phantom to demonstrate the impact on AI when modifications to imaging equipment are made.

Our study had limitations. First, a comparative analysis of different AI applications across multiple scanners was beyond the scope of this work. Likewise, a comprehensive assessment of AI across the biological variability of aneurysms and human anatomy was not our aim and should be done in patient studies. Second, due to the closed nature of the AI solution and lack of quality control interfaces, we were limited to standard AI heatmap output and cannot provide proof that missing AI responses were associated with false negative triage decisions. Third, unlike the AI, the participating neuroradiologists were able to learn from our data, which prevented us from assigning them the exact same search and detection task as the AI. Lastly, as repetitive and standardized testing in patients is precluded, we were not able to conduct the same testing in a patient cohort.

In conclusion, diagnostic AI for intracranial aneurysm detection performs inconsistently when CT image quality is not optimal and shows less resilience to data quality changes than radiologists. The AI response does not follow conventional relationships between dose, image quality, and diagnostic performance. Our findings highlight the need for standardized monitoring of AI performance, and quality control interfaces in AI solutions would facilitate this process.

## Data Availability

All data produced in the present study are available upon reasonable request to the authors.

## Acknowledgements

We thank Jelena Bevanda and Lutz Kreissl for participating in the reading experiment.

