## Supplemental Tables 1 and 2 for "Inconsistency of AI in Intracranial Aneurysm Detection with Varying Dose and Image Reconstruction"

### Supplementary Tables

Supplementary Table 1: Results of the reader assessment of aneurysm visibility. Values represent the absolute number of CT examinations in which aneurysms were rated as visible across three repeated CT examinations per dose and image reconstruction method. Means ± standard errors of the means of five readers are presented. IR = iterative reconstruction, FBP = filtered back projection, MCA = middle cerebral artery, ACoA = anterior communicating artery, BA = basilar artery, $CTDI_{vol}$ = computed tomography dose index.

| $CTDI_{vol}$ (mGy) | IR | | | FBP | | |
|---|---|---|---|---|---|---|
| | ACoA | MCA | BA | ACoA | MCA | BA |
| 0.47 | 2.2 ± 0.37 | 3 | 1.2 ± 0.5 | 2.2 ± 0.37 | 2.8 ± 0.2 | 0.8 ± 0.58 |
| 1.01 | 3 | 3 | 2.2 ± 0.49 | 2.8 ± 0.2 | 3 | 2.8 ± 0.2 |
| 2.02 | 3 | 3 | 2.8 ± 0.2 | 3 | 3 | 2.6 ± 0.25 |
| 3 | 3 | 3 | 3 | 3 | 3 | 3 |
| 4.01 | 3 | 3 | 3 | 3 | 3 | 3 |
| 5.02 | 3 | 3 | 3 | 3 | 3 | 3 |
| 6.03 | 3 | 3 | 3 | 3 | 3 | 3 |
| 7.05 | 3 | 3 | 3 | 3 | 3 | 3 |
| 8.06 | 3 | 3 | 3 | 3 | 3 | 3 |
| 9.07 | 3 | 3 | 3 | 3 | 3 | 3 |
| 10.08 | 3 | 3 | 3 | 3 | 3 | 3 |
| 11.09 | 3 | 3 | 3 | 3 | 3 | 3 |
| 12.11 | 3 | 3 | 3 | 3 | 3 | 3 |
| 13.04 | 3 | 3 | 3 | 3 | 3 | 3 |
| 14.06 | 3 | 3 | 3 | 3 | 3 | 3 |
| 15.07 | 3 | 3 | 3 | 3 | 3 | 3 |
| 16.04 | 3 | 3 | 3 | 3 | 3 | 3 |
| 17.05 | 3 | 3 | 3 | 3 | 3 | 3 |
| 18.07 | 3 | 3 | 3 | 3 | 3 | 3 |
| 19.08 | 3 | 3 | 3 | 3 | 3 | 3 |
| 20.09 | 3 | 3 | 3 | 3 | 3 | 3 |

Supplementary Table 2: Results of the reader assessment of image quality. Values represent image quality ratings per aneurysm, dose, and image reconstruction method. Means ± standard errors of the means of five readers are presented. IR = iterative reconstruction, FBP = filtered back projection, MCA = middle cerebral artery, ACoA = anterior communicating artery, BA = basilar artery, $CTDI_{vol}$ = computed tomography dose index.

| $CTDI_{vol}$ (mGy) | IR | | | FBP | | |
|---|---|---|---|---|---|---|
| | ACoA | MCA | BA | ACoA | MCA | BA |
| 0.47 | 0.87 ± 0.17 | 1.20 ± 0.11 | 0.47 ± 0.17 | 0.87 ± 0.17 | 1.00 ± 0.10 | 0.27 ± 0.12 |
| 1.01 | 1.67 ± 0.16 | 1.87 ± 0.17 | 1.27 ± 0.18 | 1.27 ± 0.18 | 1.47 ± 0.13 | 1.07 ± 0.12 |
| 2.02 | 2.00 ± 0.14 | 2.33 ± 0.13 | 1.60 ± 0.13 | 2.00 ± 0.17 | 2.27 ± 0.12 | 1.47 ± 0.24 |
| 3 | 2.40 ± 0.16 | 2.53 ± 0.13 | 2.20 ± 0.14 | 1.87 ± 0.19 | 2.27 ± 0.15 | 1.80 ± 0.22 |
| 4.01 | 2.20 ± 0.17 | 2.53 ± 0.13 | 2.20 ± 0.17 | 2.13 ± 0.19 | 2.47 ± 0.13 | 2.20 ± 0.17 |
| 5.02 | 2.60 ± 0.13 | 3 | 2.67 ± 0.13 | 2.53 ± 0.17 | 2.87 ± 0.09 | 2.47 ± 0.17 |
| 6.03 | 2.73 ± 0.12 | 2.93 ± 0.07 | 2.67 ± 0.13 | 2.73 ± 0.12 | 2.93 ± 0.07 | 2.60 ± 0.13 |
| 7.05 | 2.67 ± 0.13 | 2.87 ± 0.09 | 2.73 ± 0.12 | 2.60 ± 0.13 | 2.93 ± 0.07 | 2.60 ± 0.13 |
| 8.06 | 2.67 ± 0.13 | 2.93 ± 0.07 | 2.67 ± 0.13 | 2.60 ± 0.16 | 2.87 ± 0.09 | 2.60 ± 0.13 |

| | | | | | | |
|---|---|---|---|---|---|---|
| 9.07 | 2.67 ± 0.13 | 3 | 2.87 ± 0.09 | 2.73 ± 0.12 | 2.93 ± 0.07 | 2.73 ± 0.12 |
| 10.08 | 2.87 ± 0.09 | 3 | 2.80 ± 0.11 | 2.80 ± 0.11 | 3 | 2.87 ± 0.09 |
| 11.09 | 2.60 ± 0.16 | 2.93 ± 0.07 | 2.67 ± 0.13 | 2.73 ± 0.12 | 2.93 ± 0.07 | 2.87 ± 0.09 |
| 12.11 | 2.80 ± 0.11 | 2.93 ± 0.07 | 2.87 ± 0.09 | 2.87 ± 0.09 | 3 | 2.73 ± 0.12 |
| 13.04 | 2.80 ± 0.11 | 3 | 2.80 ± 0.11 | 2.80 ± 0.11 | 3 | 2.93 ± 0.07 |
| 14.06 | 2.80 ± 0.11 | 2.93 ± 0.07 | 3 | 2.80 ± 0.11 | 3 | 2.80 ± 0.11 |
| 15.07 | 2.67 ± 0.16 | 2.93 ± 0.07 | 2.87 ± 0.09 | 2.73 ± 0.12 | 3 | 2.80 ± 0.11 |
| 16.04 | 2.87 ± 0.09 | 3 | 2.93 ± 0.07 | 2.93 ± 0.07 | 3 | 2.93 ± 0.07 |
| 17.05 | 2.80 ± 0.14 | 3 | 2.93 ± 0.07 | 2.73 ± 0.12 | 2.87 ± 0.09 | 2.67 ± 0.13 |
| 18.07 | 2.80 ± 0.11 | 3 | 2.93 ± 0.07 | 2.80 ± 0.11 | 3 | 2.93 ± 0.07 |
| 19.08 | 2.87 ± 0.09 | 3 | 2.80 ± 0.11 | 2.87 ± 0.09 | 3 | 2.87 ± 0.09 |
| 20.09 | 2.87 ± 0.09 | 3 | 3 | 2.67 ± 0.13 | 2.93 ± 0.07 | 2.73 ± 0.12 |
